# Three-dimensional cranio-facial landmark detection in CT slices from a publicly available database, using multi-phased regression networks on a personal computer

**DOI:** 10.1101/2021.03.21.21253999

**Authors:** Soh Nishimoto, Takuya Saito, Hisako Ishise, Toshihiro Fujiwara, Kenichiro Kawai, Maso Kakibuchi

**Author notes:** Contact: Soh Nishimoto, Department of Plastic Surgery, Hyogo College of Medicine., 1-1 Mukogawa-cho, Nishinomiya, Hyogo, Japan. 663-8131.

## Abstract

**Aim:** Geometrical assessments to comprehend the shape of an object are done based on characteristic landmarks. Computer assisted tomography (CT) images, horizontal slices as two-dimensional pictures, can be digitally restructured into virtual three-dimensional objects. Automatic detection of the landmarks, if developed, will be a great help not only medically, but also for anthropologically. The aim of this study is to develop an automated system to predict three-dimensional coordinate values of cranio-facial landmarks in sequences of CT slices.

**Methods:** CT images were obtained from a publicly available database. Digital reconstruction was done to obtain three dimensional models. Sixteen landmarks were plotted on the models and coordinate values of them were recorded. Multi-phased deep learning system was constructed. For the first phase, 512 x 512 pixels images were resized to 96 x 96 pixels. A regression deep learning network was trained with 90 training data. For the second phase, for each landmark, 100 x 100 pixels images were cropped from the original images. Sixteen models were trained. For the third phase, 50 x 50 pixels images were cropped, and models were trained.

**Results:** Three-dimensional error for the first phase, testing 30 data, was 11.60 pixels in average. (1 pixel = 500 / 512 mm) For the second phase, it was significantly improved to 4.66 pixels. For the third phase, it was significantly progressed to 2.89. This was comparable to the gaps between the landmarks, plotted by two experienced practitioners.

**Discussion:** The calculation volume required to process three-dimensional pile of images is tremendous. One solution may be to compress the images, but detailed information may be lost during the process. Our proposing method of multi-phased prediction, coarse detection first and narrowing down the detection area, may be a possible solution, within the physical limitation of memory and computation.

## 1. Introduction

Measuring distances between characteristic landmarks and angles between certain planes determined by the points, is a useful approach to comprehend the shape of an object. Cephalometry, first introduced by Broadbent[Broadbent, 1931] and Hofrath[Hofrath, 1931] in 1931, has been and still is one of the most helpful modalities in evaluating cranio-maxillo-facial configurations. It is to evaluate two-dimensional X-ray images, taken with stabilization of the examinee’s head in a standard position. Geometrical assessments are done based on the anatomical landmarks. Inherently, this method encompasses some kinds of inaccuracy and incapability, because it is to evaluate objects in two-dimensionally projected images, which are originally in three dimensions. Computer assisted tomography (CT) has become popular in daily clinical practice. CT images, horizontal slices as two-dimensional pictures, are usually stored in DICOM (Digital Imaging and Communications in Medicine) formatted files. They can be digitally restructured into virtual three-dimensional objects. 3D printing can also be done. They visually help people to comprehend the bodies. Three-dimensional measurement based on the anatomical landmarks can be done on the objects. In orthodontic field, three-dimensional cephalogram is becoming popular. Locating anatomical landmarks demands time and expertise. Automatic detection of the landmarks, if developed, will be a great help not only medically, but also for anthropologically.

Authors previously developed an automated system to predict landmarks on the two-dimensional cephalogram images[Nishimoto et al., 2019][Nishimoto, 2020][Nishimoto et al., 2020]. Utilizing multi-phased regression deep learning neural networks enhanced the prediction accuracy[Nishimoto, 2020][Nishimoto et al., 2020][Kim et al., 2020]. In this study, the same approach was attempted to predict three-dimensional coordinate values of landmarks in sequences of CT images.

## 2. Materials and Methods

### 2.1 Personal computer

All procedures were done on a desk-top personal computer: CPU (Central Processing Unit): AMD Ryzen 7 2700X 3.70GHz (Advanced Micro Systems, Sunnyvale, CA, USA), memory: 64.0GB, GPU: GeForce RTX2080 8.0GB ((nVIDIA, Santa Clara, CA, USA), Windows 10 pro (Microsoft Corporations, Redmond, WA, USA). Python 3.7 (Python Software Foundation, DE USA): a programing language, was used under Anaconda 15 (FedoraProject. http://fedoraproject.org/wiki/Anaconda#Anaconda_Team_Emeritus) as an installing system, and Spyder 4.1.4 as an integrated development environment. Keras 2.31 (https://keras.io/): the deep learning library, written in Python was run on TensorFlow 1.14.0 (Google, Mountain View, CA, USA). GPU computation was employed through CUDA 10.0 (nVIDIA). For 3D reconstruction, slicer 4.11 (www.slicer.org) was used with Jupyter Notebook (https://jupyter.org/). OpenCV 3.1.0 libraries (https://docs.opencv.org/3.1.0/) were used in image processing.

### 2.2 Datasets

#### (1) CT images

From The Cancer Imaging Archive Public Access (wiki.cancerimagingarchive.net), Head-Neck-Radiomics-HN1[Blake, 2020], the collection of CT images from head and neck squamous cell carcinoma patients was retrieved. It consists of the folder of each patient, containing 512 x 512 pixels DICOM images (0 to 3000 for each pixel) taken at 5 mm intervals in the cephalo-caudal direction. The order of the images was checked and images from the top of the head to the mandible were extracted for 120 cases. The largest number of extracted images for a patient was 81. As a calibration marker, a 512-pixel length and width cross were added to the most caudal images (Supple.1).

#### (2) 3D reconstruction (STL file creation)

DICOM CT image sequence for each case was processed with 3D slicer kernel using Jupyter notebooks. With a python script process[Lasso, 2020], bony parts were segmented and reconstructed into 3D images and stored as STL files (Supple.2).

#### (3) Plotting anatomical landmarks

Each STL file was imported into blender and three-dimensionally expanded by 512 pixels / 500 mm (Supple.3). Spheres with 1 pixel radius were placed as the landmarks.

The landmarks are listed in Table 1. and shown in Figure 1. Three-dimensional coordinate values (x, y, z) of the imported STL and spheres were obtained and exported as an array of 120 cases x 16 points x 3 (Supple.4). Two practitioners, with 31 and 10 year-experience, respectively plotted landmarks. The coordinate values plotted by the senior was used as the ground zero (Supple.5).

**Table 1.**
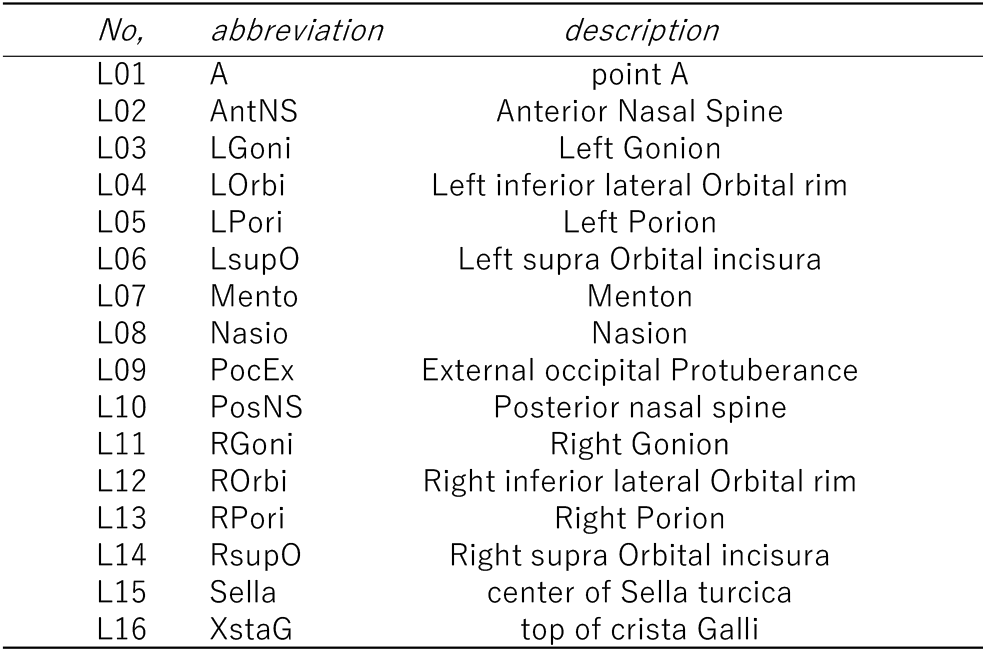

| No, | abbreviation | description |
| --- | --- | --- |
| L01 | A | point A |
| L02 | AntNS | Anterior Nasal Spine |
| L03 | LGoni | Left Gonion |
| L04 | LOrbi | Left inferior lateral Orbital rim |
| L05 | LPori | Left Porion |
| L06 | LsupO | Left supra Orbital incisura |
| L07 | Mento | Menton |
| L08 | Nasio | Nasion |
| L09 | PocEx | External occipital Protuberance |
| L10 | PosNS | Posterior nasal spine |
| L11 | RGoni | Right Gonion |
| L12 | ROrbi | Right inferior lateral Orbital rim |
| L13 | RPori | Right Porion |
| L14 | RsupO | Right supra Orbital incisura |
| L15 | Sella | center of Sella turcica |
| L16 | XstaG | top of crista Galli |

**Figure 1.**
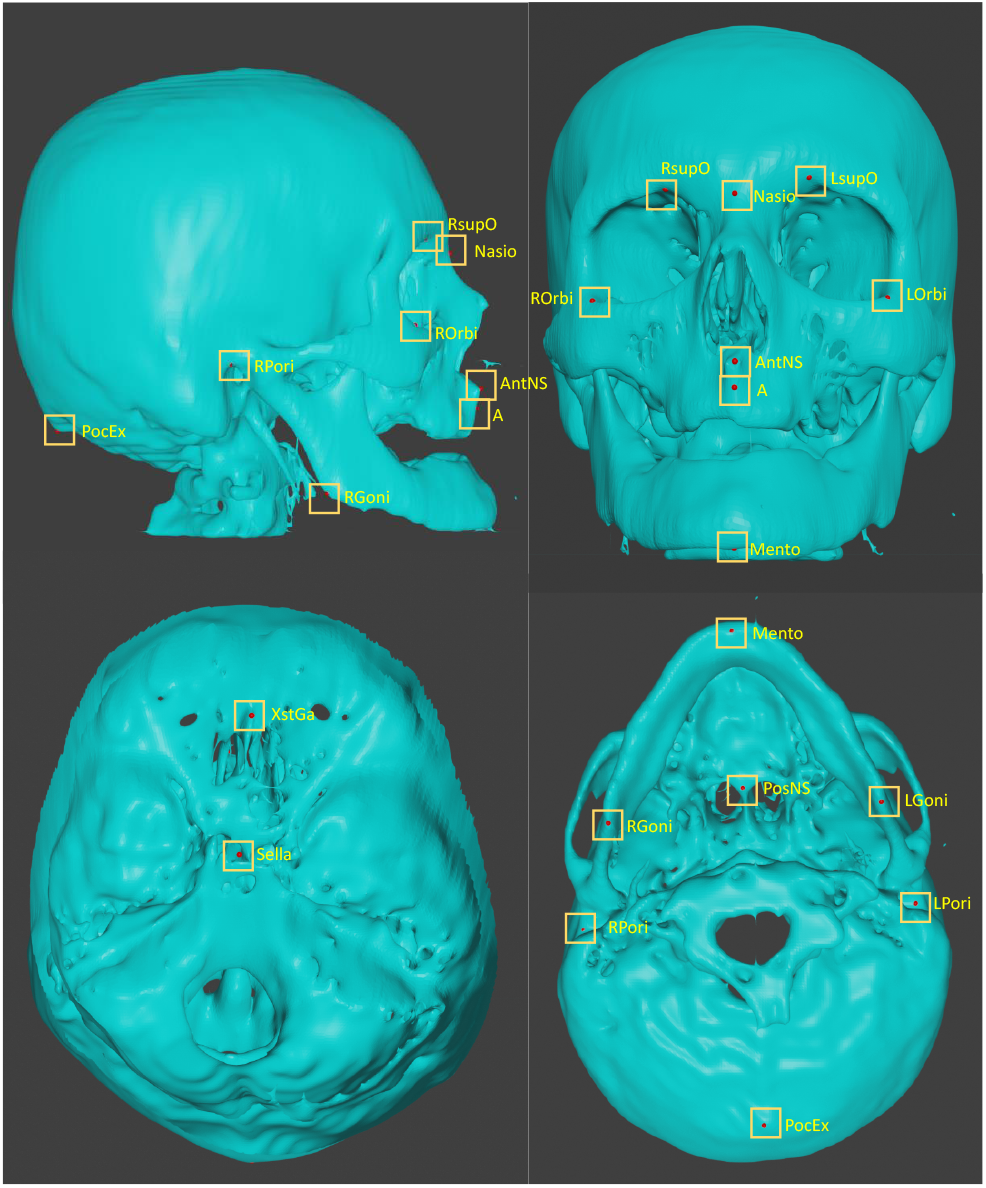
Three dimensionally plotted landmarks

#### (4) Neural Networks and Learning

##### [1] 1st phase deep learning (Figure 2.)

With OpenCV, each CT image (512 x 512 pixels DICOM image) was binarized to segment bone with 1100 as threshold and compressed to 96 x 96 pixels. For each case, they were stacked up from the bottom to form a three-dimensional array of 96 x 96 x 81 (Supple.6,7). As training data, 90 cases were assigned, and 30 were as testing data.

**Figure 2.**
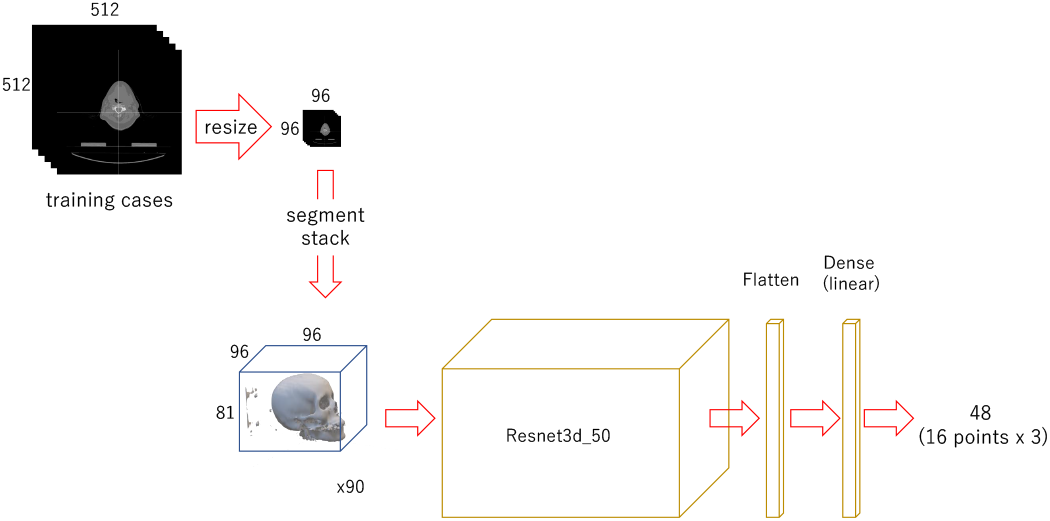
Diagram of the 1st phase deep learning

A regression deep learning model, modified (only the last activation layer was changed from “softmax” to “linear”) Resnet 3d-50 [Jihong, 2019], built with 96 x 96 x 81 as input and 48 as output (Supple.8). It was trained for 150 epochs.

##### [2] 2nd phase deep learning (Figure 3.)

A 100 x 100 pixel image was cropped out from each original image with OpenCV, centered on the x and y coordinates of each landmark, and piled up from the bottom to form a 100 x 100 x 81 3D array. The images were also cropped out at shifted positions in the x and y directions and stacked in the same way to obtain the positions of feature points in each array (3240 sets in total)(Supple.9). For each landmark, the modified Resnet 3d-50 model for regression with 100 x 100 x 81 as input and 3 as output (Supple.10) was trained for 100 epochs.

**Figure 3.**
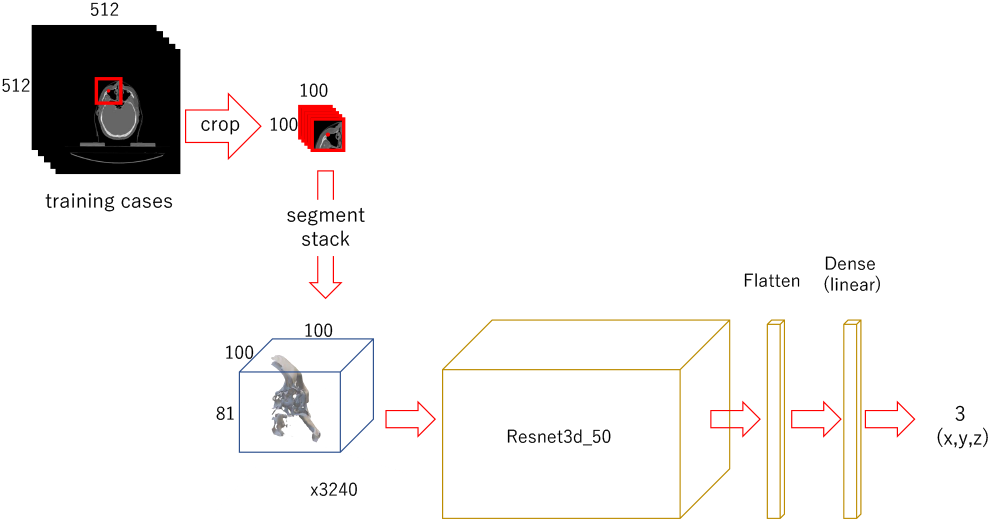
Diagram of the 2nd phase deep learning

##### 3rd phase deep learning (Figure 4.)

A 50 x 50 pixel image was cropped out from each original image, in the same way for the 2nd phase (Supple.11) Stacks of 50 x 50 x 81 were obtained. For training, 3240 sets of data for each landmark were used. Modified Resnet 3d-50 with input of 50 x 50 x 81 and 3 as output (Supple.12) was trained for 150 epochs.

**Figure 4.**
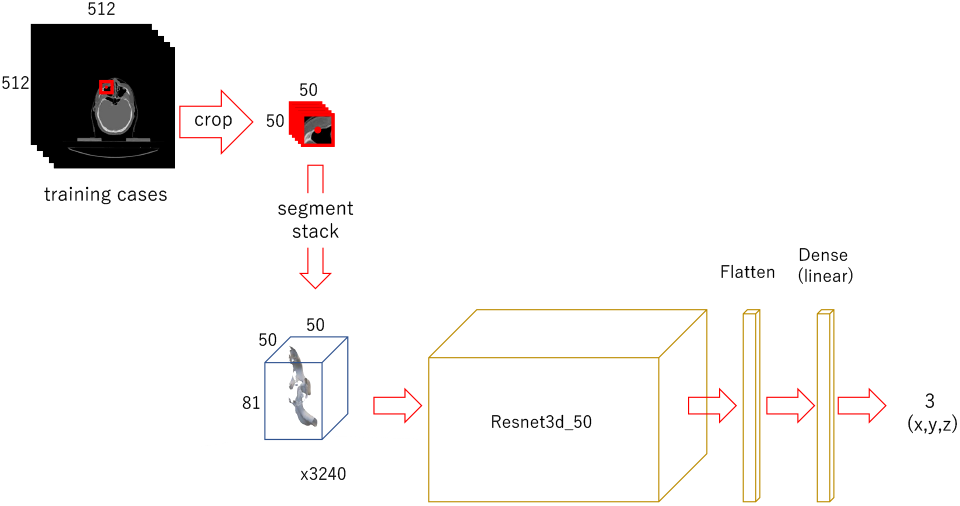
Diagram of the 3rd phase deep learning

#### (5) Evaluation (Figure 5.)

For evaluation, 30 examples that were not used in the training were employed.

##### [1] 1st phase prediction

The 96 x 96 x 81 3D array of the 30 cases for validation was fed to the trained 1st phase model to predict the 3D coordinates of feature points.

**Figure 5.**
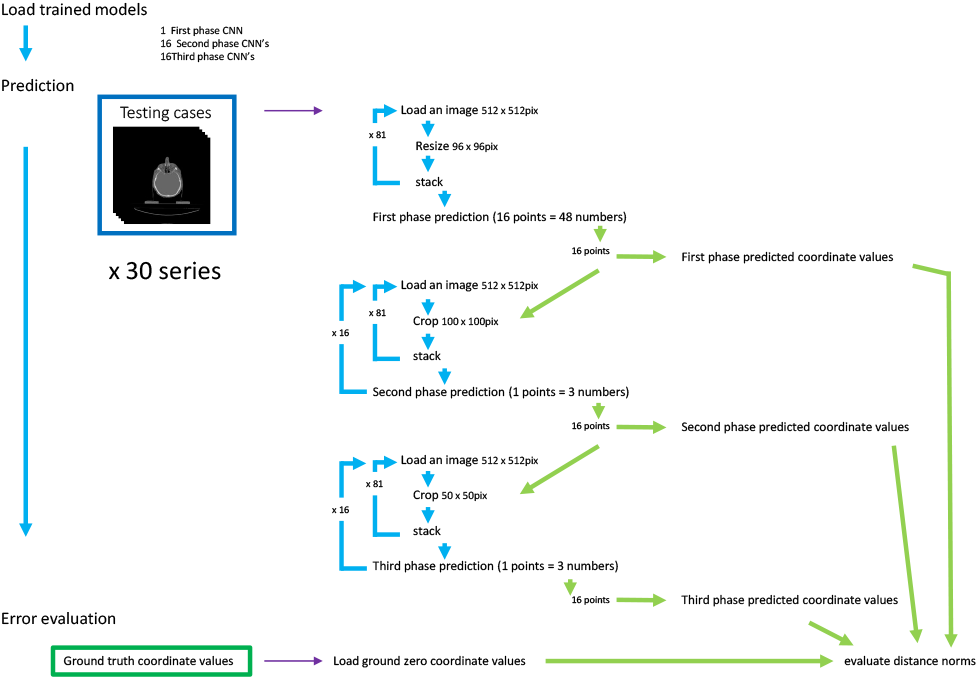
Prediction and evaluation

##### [2] 2nd phase prediction

The 100 x 100 pixel images were cropped from the original validation images, centered on each of the 16 coordinates obtained in the 1st phase prediction, and piled up into 100 x 100 x 81 3D arrays. They were used to predict the coordinates of each feature point with the trained 2nd phase models.

##### [3] 3rd phase prediction

For each landmark, 50 x 50 pixel images were cropped, centered on each of the coordinates obtained in the 2nd phase prediction. They were stacked up to 50 x 50 x 81 arrays and fed to the respective 16 trained 3rd phase models.

##### [4] Prediction error evaluation

The distances between the predicted coordinates and the manually plotted ground truth coordinates were calculated as the absolute value in the x, y, and z directions. The square root of the sum of the squares of each was used as the 3D distance.

##### [5] Inter-observer gap

The distances between the landmarks plotted by the senior and the junior practitioner were calculated as the inter-observer gap.

##### [6] Statistic analysis

Multiple comparisons were done using scikit-posthocs(@rola_satoru).

## 3. Results

### 3.1 1st phase prediction error

Overall average three-dimensional distance between the predicted points and the ground zero was 11.60 pixels (1 pixel = 500 / 512 mm) (Table 2.). Per landmark prediction errors are shown in Figure 6. Between axis directions, error for x-axis was significantly smaller than the others, and y-axis was the largest (Figure 7.).

**Table 2.** Three-dimentional prediction errors in 480 landmarks of testing data (pixel = 500/512 mm)

|  | 1st phase | 2nd phase | 3rd phase | inter-observer gaps |
| --- | --- | --- | --- | --- |
| *average* | 11.6 | 4.66 | 2.88 | 3.08 |
| *median* | 10.89 | 4.22 | 2.56 | 2.40 |
| *stdev* | 5.64 | 2.27 | 1.67 | 2.64 |

\* p<0.001 by conover test

**Figure 6.**
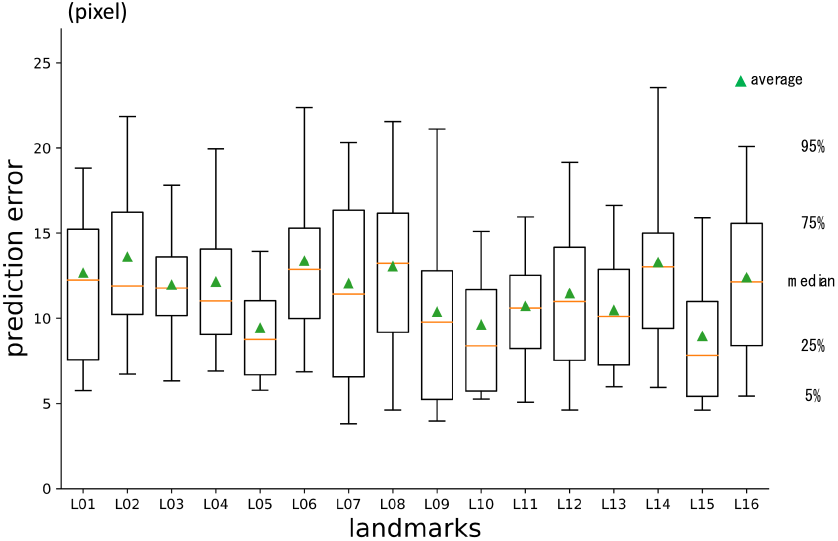
1st phase prediction errors per landmark (pixel = 500 / 512 mm)

**Figure 7.**
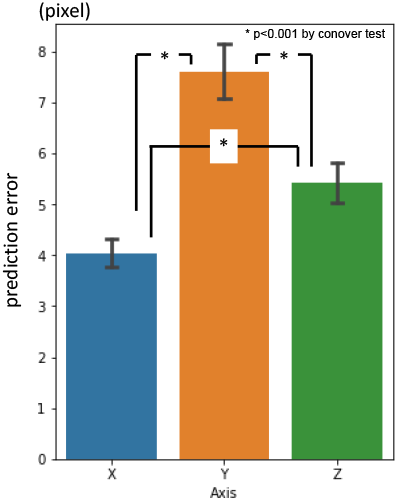
1st phase prediction errors per axis (pixel = 500 / 512 mm)

### 3.2 2nd phase prediction error

In average, three-dimensional prediction errors was 4.66 pixels (Table 2.). It was significantly smaller than 1st phase prediction. Per landmark errors are shown in Figure 8. Error for y-axis direction was larger than the others. Error for z-axis was the smallest (Figure 9.).

**Figure 8.**
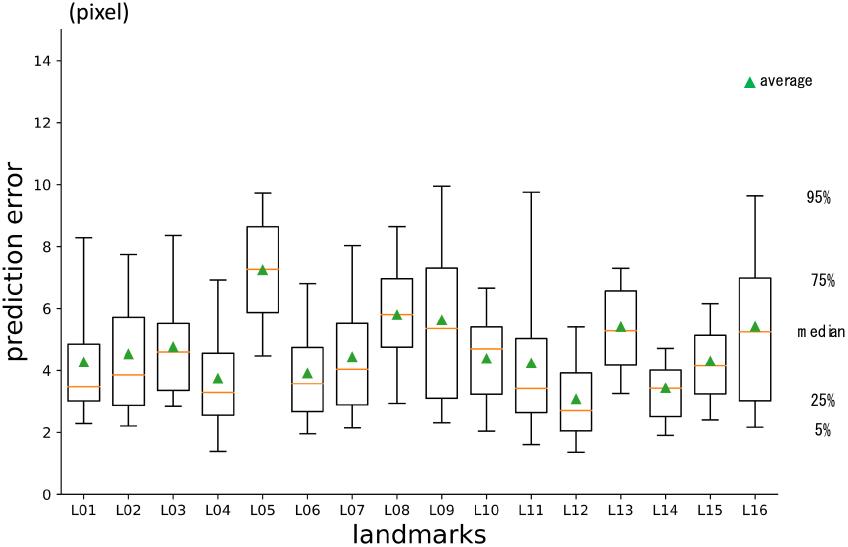
2nd phase prediction errors per landmark (pixel = 500 / 512 mm)

**Figure 9.**
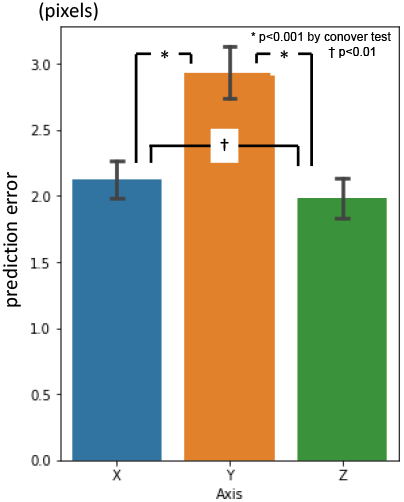
2nd phase prediction errors per axis (pixel = 500/ 512 mm)

### 3.3 3rd phase prediction error

Three-dimensional prediction error was 2.88 pixels in average (Table 2.). It was significantly smaller than 2nd phase prediction. Errors per landmark are shown in Figure 10. There was no significant difference between axes (Figure 11.).

**Figure 10.**
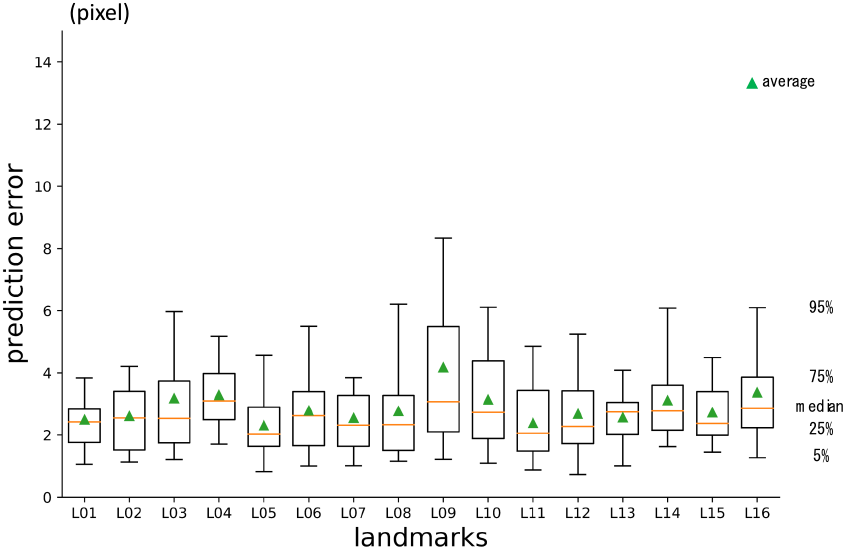
3rd phase prediction errors per landmark (pixel= 500 / 512 mm)

**Figure 11.**
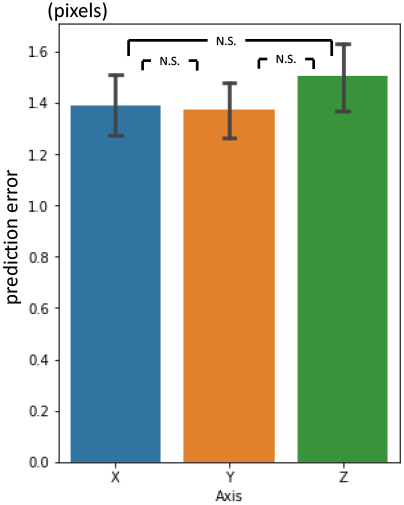
3rd phase prediction errors per axis (pixel = 500/ 512 mm)

### 3.4 Inter-phase and inter-observer plotting gap

There were significant differences between prediction errors by phases. Inter-observer plotting gap was 3.08 pixels in average, 2.40 as median with standard deviation of 2.64 in total 120 data. The 3rd phase prediction error for 30 testing data was the same level as the inter-observer plotting gaps (Table 2.).

## 4. Discussion

### 4.1 3D landmark detection

The history of standard cephalometry, since Broadbent[Broadbent, 1931] and Hofrath[Hofrath, 1931] first proposed. is quite long. There have been many reports on automatic landmark detection systems for cephalograms[Forsyth and Davis, 1996][Giordano et al., 2005][Tanikawa et al., 2009][Wang et al., 2016][Lindner et al., 2016][Arik et al., 2017][Nishimoto et al., 2019][Nishimoto et al., 2020], which are in two-dimensions. Rebuilding 3D models from series of two-dimensional CT images became possible. In comparison with 2D cephalograms, reports on automatic landmark detection systems for 3D images are small in number[Shahidi et al., 2014] [Dot et al., 2020].

Some previous automated 3D landmark detectors in craniofacial area employed knowledge-based methods[Mestiri and Kamel, 2014][Gupta et al., 2015][Codari et al., 2017]. This approach requires knowledge building procedure, which is customized by landmark to landmark. Deep learning is one of the most emerging techniques in machine-learning. It is categorized as a supervised learning that to find out rules between input and output of training datasets. All we need is to prepare the data, and the machine will figure out the laws. It is very versatile. Authors previously developed automatic systems to detect landmarks in 2D cephalograms using deep learning[Nishimoto et al., 2020][Nishimoto et al., 2019][Nishimoto, 2020].

### 4.2 Multi-phased deep learning

The processing speed of computers and the amount of memory they installed have increased at a dizzying rate. Though, the amount of computation, required to process images with deep learning is enormous. The calculation volume required to process three-dimensional piles of images, based on spatial or time axes, is incomparable to that required for two-dimensional images. One solution is to compress the images[Kang et al., 2018] and input them to deep learning, but the compression process results in the loss of detailed information. In this study, we applied the method of multi-phase deep learning, used to predict landmarks in 2D cephalograms[Nishimoto, 2020][Nishimoto et al., 2020], to 3D craniofacial images. Prediction errors became smaller as the phase advanced. Coarse detection was done with the first phase model and father narrowing down was done based on the prediction of the previous phase. The prediction errors by the 3rd phase prediction were comparable to the gaps between the experienced practitioners.

This study was conducted on a personal computer. Within the physical limitation of memory and computation, multi-phase deep learning may be a solution to deal with large-scaled images.

There could find two reports in 3D landmark detection using fully convolutional neural networks (FCN) with high precision[Torosdagli et al., 2019][O’Neil et al., 2018]. But in general, FCN is computationally expensive and slow.

Our system is very simple. Through all three phases, the main part of the models used were the same Resnet 3d-50 modified for regression. There may be ways to design the system in end-to-end fashion. Again, within the calculation limit, this sequential system was practical for the authors.

### 4.3 Database

In the previous studies[Nishimoto et al., 2020][Nishimoto, 2020][Nishimoto, 2020] of 2D cephalograms, there was a database published at ISBI 2015 [Wang et al., 2016] and a prior benchmark using it[Arik et al., 2017]. Authors were unable to obtain a database of feature point coordinates for craniofacial CT. In the current status in Japan, it is not easy to access and build database of patient information, even for clinicians as the authors. Hence, authors constructed a database from publicly available image sets[Blake, 2020]. Therefore, the prediction accuracy could not be compared with the other methods. The image sets used were from patients with head and neck tumors. There was no information on age or genders. Most of the images were probably from elderly patients, and there were many missing teeth. Many of them were in an open bite position,

### 4.4 Three-dimensional plot

The plotting featured points one by one took a long time. Plotting anatomical feature points in 3D requires multiple perspectives. In this study, we used five viewpoints in blender. In addition, it is often necessary to pan, zoom in and out, Bone ridges are formed by curves, not by sharp angles, so it was difficult and sometimes impossible to plot them accurately. To maintain consistency as much as possible, one feature point was plotted for all cases in succession. Standard deviations of the coordinate values were 33.85 for x axis, 48.68 for y axis and 43.08 for z axis, including both training and testing data. The differences may have affected the 1st phase prediction accuracy per axis (Figure 7.).

### 4.5 CT slice thickness

The craniofacial CT data used in this study were taken at 5 mm intervals in the cephalo-caudal direction. The maximum number of slices in the z-axis direction was 81. No compression in the z-axis direction was performed as our machine was capable to deal the image piles with 81 depth.

In clinical practice, slices of less than 1 mm are commonly used to obtain detailed bone information (so-called thin slices). To apply these images in real clinical situations, such as navigation systems used in surgery, it is necessary to support these images.

Cone-beam computed tomography (CBCT), which has been becoming popular in orthodontists and otolaryngologists, can get images with small voxels. When based on the detailed images, highly accurate estimation can be expected. Though, the computation volume will increase as the information to be processed increase. Our proposing method of multi-phased prediction, coarse detection first and narrowing down the detection area, may be a possible solution.

## Supporting information

Supple 1. a sample of bottom image

Supple 2. Slicer Jupyter notebook to make STLs

Supple 3. an example of blender file

Supple 4. blender python script to collect coordinate values

Supple 5. pkl file for coordinate values: filenames,pointnames,X,Y,Z= pickle.load(f)

Supple 6. python code to process dicom files to numpy

Supple 7.pkl file of compressed 3Dimages: dda = pickle.load(f) (120,81,96,96)

Supple 8. python code to train 1st phase

Supple 9. python code to crop images for 2nd phase

Supple10. python code to train 2nd phase

Supple 11. python code to crop images for 3rd phase

Supple 12. python code to train 3rd phase

## Data Availability

Original CT data was obtained from a publicly available database: The Cancer Imaging Archive Public Access (wiki.cancerimagingarchive.net), Head-Neck-Radiomics-HN1

https://wiki.cancerimagingarchive.net/display/Public/Head-Neck-Radiomics-HN1

## 5. Acknowledgement

This study was partly supported by Nakatani Foundation Research grant.

## Notes

### Competing Interest Statement

The authors have declared no competing interest.

## References

1. @rola_satoru. Pythonで統計検定(多重検定)?: Scikit_posthocs - Qiita. 2020, https://qiita.com/rola_satorus: A systematic re 672891f80.

2. Arik Sercan Ö., et al. “Fully Automated Quantitative Cephalometry Using Convolutional Neural Networks.” Journal of Medical Imaging, vol. 4, no. 1, SPIE-Intl Soc Optical Eng, Jan. 2017, p. 014501, doi:10.1117/1.jmi.4.1.014501.

3. Blake Geri. Head-Neck-Radiomics-HN1 - The Cancer Imaging Archive (TCIA) Public Access - Cancer Imaging Archive Wiki. 2020, https://wiki.cancerimagingarchive.net/display/Public/Head-Neck-Radiomics-HN1.

4. Broadbent B Holly. “A New X-Ray Technique and Its Application to Orthodontia.” The Angle Orthodontist:, vol. 1, no. 2, 1931, pp. 45–66, doi:10.1043/0003-3219(1981)051<0093:ANXTAI>2.0.CO;2.

5. Codari Marina., et al. “Computer-Aided Cephalometric Landmark Annotation for CBCT Data.” International Journal of Computer Assisted Radiology and Surgery, vol. 12, no. 1, Springer Verlag, Jan. 2017, pp. 113–21, doi:10.1007/s11548-016-1453-9.

6. Dot Gauthier., et al. “Accuracy and Reliability of Automatic Three-Dimensional Cephalometric Landmarking.” International Journal of Oral and Maxillofacial Surgery, vol. 49, no. 10, Churchill Livingstone, 1 Oct. 2020, pp. 1367–78, doi:10.1016/j.ijom.2020.02.015.

7. Forsyth D B., and Davis D N. “Assessment of an Automated Cephalometric Analysis System.” European Journal of Orthodontics, vol. 18, no. 5, Oxford University Press, 1996, pp. 471–78, doi:10.1093/ejo/18.5.471.

8. Giordano Daniela., et al. “Automatic Landmarking of Cephalograms by Cellular Neural Networks.” Artificial Intelligence in Medicine. AIME 2005. Lecture Notes in Computer Science, Springer, Berlin, Heidelberg, 2005, pp. 333–42, doi:10.1007/11527770_46.

9. Gupta Abhishek., et al. “A Knowledge-Based Algorithm for Automatic Detection of Cephalometric Landmarks on CBCT Images.” International Journal of Computer Assisted Radiology and Surgery, vol. 10, no. 11, Springer Verlag, Nov. 2015, pp. 1737–52, doi:10.1007/s11548-015-1173-6.

10. Hofrath H. “Die Bedeutung Der Roentgenfern Der Kiefer Anomalien.” Fortschr Orthodontic, vol. 1, 1931, pp. 232–48.

11. Jihong Ju. GitHub - JihongJu/Keras-Resnet3d: Implementations of ResNets for Volumetric Data, Including a Vanilla Resnet in 3D. 2019, https://github.com/JihongJu/keras-resnet3d.

12. Kang Sung Ho., et al. Automatic Three-Dimensional Cephalometric Annotation System Using Three-Dimensional Convolutional Neural Networks. Nov. 2018, http://arxiv.org/abs/1811.07889.

13. Kim Hannah., et al. “Web-Based Fully Automated Cephalometric Analysis by Deep Learning.” Computer Methods and Programs in Biomedicine, vol. 194, Elsevier Ireland Ltd, Oct. 2020, p. 105513, doi:10.1016/j.cmpb.2020.105513.

14. Lasso Andras. Lassoan/ExtractSkin.Py. 2020, https://gist.github.com/lassoan/1673b25d8e7913cbc245b4f09ed853f9.

15. Lindner Claudia., et al. “Fully Automatic System for Accurate Localisation and Analysis of Cephalometric Landmarks in Lateral Cephalograms.” Scientific Reports, vol. 6, Nature Publishing Group, Sept. 2016, p. 33581, doi:10.1038/srep33581.

16. Mestiri Makram. and Hamrouni Kamel. “Reeb Graph for Automatic 3D Cephalometry.” International Journal of Image Processing, vol. 8, 2014, pp. 2014–31.

17. Nishimoto Soh., et al. “Personal Computer-Based Cephalometric Landmark Detection with Deep Learning, Using Cephalograms on the Internet.” Journal of Craniofacial Surgery, vol. 30, no. 1, 2019, doi:10.1097/SCS.0000000000004901.

18. Nishimoto Soh. Cephalometric Landmark Location with Multi-Phase Deep Learning. ?般社団法人人工知能学会, 2020, doi:10.11517/PJSAI.JSAI2020.0_2Q6GS1001.

19. Nishimoto Soh. et al.. “Locating Cephalometric Landmarks with Multi-Phase Deep Learning.” MedRxiv, medRxiv, 14 July 2020, p. 2020.07.12.20150433, doi:10.1101/2020.07.12.20150433.

20. Nishimoto Soh. et al.. “Personal Computer-Based Cephalometric Landmark Detection with Deep Learning, Using Cephalograms on the Internet.” Journal of Craniofacial Surgery, vol. 30, no. 1, Lippincott Williams and Wilkins, Jan. 2019, pp. 91–95, doi:10.1097/SCS.0000000000004901.

21. O’Neil Alison Q., et al. “Attaining Human-Level Performance with Atlas Location Autocontext for Anatomical Landmark Detection in 3D CT Data.” Lecture Notes in Computer Science (Including Subseries Lecture Notes in Artificial Intelligence and Lecture Notes in Bioinformatics), vol. 11131 LNCS, Springer Verlag, May 2018, pp. 470–84, http://arxiv.org/abs/1805.08687.

22. Shahidi Shoaleh., et al. “The Accuracy of a Designed Software for Automated Localization of Craniofacial Landmarks on CBCT Images.” BMC Medical Imaging, vol. 14, no. 1, BioMed Central Ltd., Sept. 2014, doi:10.1186/1471-2342-14-32.

23. Tanikawa Chihiro., et al. “Automated Cephalometry: System Performance Reliability Using Landmark-Dependent Criteria.” Angle Orthodontist, vol. 79, no. 6, Allen Press, Nov. 2009, pp. 1037–46, doi:10.2319/092908-508R.1.

24. Torosdagli Neslisah., et al. “Deep Geodesic Learning for Segmentation and Anatomical Landmarking.” IEEE Transactions on Medical Imaging, vol. 38, no. 4, Institute of Electrical and Electronics Engineers Inc., Apr. 2019, pp. 919–31, doi:10.1109/TMI.2018.2875814.

25. Wang Ching Wei., et al. “A Benchmark for Comparison of Dental Radiography Analysis Algorithms.” Medical Image Analysis, vol. 31, Elsevier B.V., July 2016, pp. 63–76, doi:10.1016/j.media.2016.02.004.

