## Supplementary figures and images for "Three-dimensional cranio-facial landmark detection in CT slices from a publicly available database, using multi-phased regression networks on a personal computer"

### Supple 1. a sample of bottom image

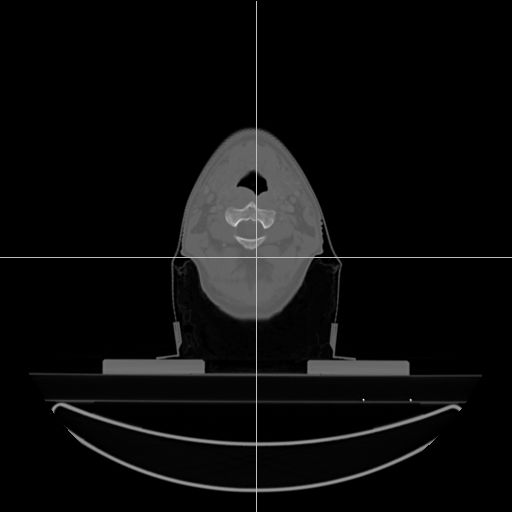
