## Supplementary material for "Three-dimensional cranio-facial landmark detection in CT slices from a publicly available database, using multi-phased regression networks on a personal computer": Supple 2. Slicer Jupyter notebook to make STLs: S2DICOMCTtoSTLwithSlicer.ipynb

```
# %% [code]
"""
modification of
Lasso A. lassoan/ExtractSkin.py. https://gist.github.com/lassoan/1673b25d8e7913cbc245b4f09ed853f9 
"""
import os
import DICOMLib

path='E:/CTskull/NBIA2'      #directory for CT sequences
wpath='E:/CTskull/NBIAs'     #directory to write STL files
os.chdir(path)
ddir = os.listdir(path)
ndir=len(ddir)
for sdir in ddir:
    dicomDataDir =path+'/'+sdir
    print(dicomDataDir)
    stlfile=wpath+'/'+sdir+'.stl'
    DICOMLib.importDicom(dicomDataDir)
    dicomFiles = slicer.util.getFilesInDirectory(dicomDataDir)
    loadablesByPlugin, loadEnabled = DICOMLib.getLoadablesFromFileLists([dicomFiles])
    loadedNodeIDs = DICOMLib.loadLoadables(loadablesByPlugin)
    for loadedNodeID in loadedNodeIDs:
        masterVolumeNode = slicer.mrmlScene.GetNodeByID(loadedNodeID) 

    # Create segmentation
    segmentationNode = slicer.mrmlScene.AddNewNodeByClass("vtkMRMLSegmentationNode")
    segmentationNode.CreateDefaultDisplayNodes() # only needed for display
    segmentationNode.SetReferenceImageGeometryParameterFromVolumeNode(masterVolumeNode)
    addedSegmentID = segmentationNode.GetSegmentation().AddEmptySegment("bone")

    # Create segment editor to get access to effects
    segmentEditorWidget = slicer.qMRMLSegmentEditorWidget()
    segmentEditorWidget.setMRMLScene(slicer.mrmlScene)
    segmentEditorNode = slicer.mrmlScene.AddNewNodeByClass("vtkMRMLSegmentEditorNode")
    segmentEditorWidget.setMRMLSegmentEditorNode(segmentEditorNode)
    segmentEditorWidget.setSegmentationNode(segmentationNode)
    segmentEditorWidget.setMasterVolumeNode(masterVolumeNode)

    # Thresholding
    segmentEditorWidget.setActiveEffectByName("Threshold")
    effect = segmentEditorWidget.activeEffect()
    effect.setParameter("MinimumThreshold","130")        #segmentation value for bone
    effect.setParameter("MaximumThreshold","3000")
    effect.self().onApply()
    # Make segmentation results visible in 3D
    segmentationNode.CreateClosedSurfaceRepresentation()

    # Make sure surface mesh cells are consistently oriented

    surfaceMesh = segmentationNode.GetClosedSurfaceInternalRepresentation(addedSegmentID)
    normals = vtk.vtkPolyDataNormals()
    normals.AutoOrientNormalsOn()
    normals.ConsistencyOn()
    normals.SetInputData(surfaceMesh)
    normals.Update()
    surfaceMesh = normals.GetOutput()
    # Write to STL file
    writer = vtk.vtkSTLWriter()
    writer.SetInputData(surfaceMesh)
    writer.SetFileName(stlfile)
    writer.Update()
```
