## Supplementary material for "Three-dimensional cranio-facial landmark detection in CT slices from a publicly available database, using multi-phased regression networks on a personal computer": Supple 4. blender python script to collect coordinate values

import bpy
import os,glob
import numpy as np
import csv
import pickle
cvx=[]
cvy=[]
cvz=[]
cvn=[]
def delete_object_all():
    
    for item in bpy.context.scene.collection.objects:
        bpy.context.scene.collection.objects.unlink(item)  
    for item in bpy.data.objects:
        bpy.data.objects.remove(item)  
    for item in bpy.data.meshes:
        bpy.data.meshes.remove(item)
    for item in bpy.data.materials:
        bpy.data.materials.remove(item)
    return

delete_object_all()

path="E:/CTskull/REDpoints"   #dir of skull blenders
os.chdir(path)
files = glob.glob("*")

for fn in files:
    file_path =path+'/'+fn
    inner_path = 'Object'
    with bpy.data.libraries.load(file_path) as (data_from, data_to):
        data_to.objects = [name for name in data_from.objects]
        print('These are the objs: ', data_to.objects)
    for i in range(len(data_to.objects)):
        object_name =data_to.objects[i].name
        bpy.ops.wm.append(
        filepath=os.path.join(file_path, inner_path, object_name),
        directory=os.path.join(file_path, inner_path),
        filename=object_name
        )
    for item in bpy.data.objects:
        item.name=item.name[:5]
        print(item.name)
        
    for item in bpy.data.cameras:
        bpy.data.cameras.remove(item)
    for item in bpy.data.lights:
        bpy.data.lights.remove(item)    
    
    lx = [] # list of objects x locations
    ly = [] # list of objects y locations
    lz = [] # list of objects z locations
    lun = []
    
    for collection in bpy.data.collections:
        print(collection.name)
        for obj in collection.all_objects:

    
               lun.append(obj.name)
               lx.append(obj.location.x)
               ly.append(obj.location.y)
               lz.append(obj.location.z)
               
    cvx.append(lx)
    cvy.append(ly)
    cvz.append(lz)
    cvn.append(lun)
    
    print(len(lx))
   
    
    delete_object_all()

cvx=np.asarray(cvx)
cvy=np.asarray(cvy)
cvz=np.asarray(cvz)
cvn=np.asarray(cvn)

X=(cvx,cvy,cvz,cvn)
sfile='points.pkl'              #file for coordinate values
with open(sfile, mode='wb') as f:    
    pickle.dump(X, f, protocol=4)


print (cvx.shape)
